# Antibody avidity maturation favors SARS-CoV-2 convalescents over vaccinated individuals granting breadth in neutralizability and tolerance against variants

**DOI:** 10.1101/2022.09.19.22280078

**Authors:** Yu Nakagama, Katherine Candray, Natsuko Kaku, Yuko Komase, Maria-Virginia Rodriguez-Funes, Rhina Dominguez, Tomoya Tsuchida, Hiroyuki Kunishima, Etsuko Nagai, Eisuke Adachi, Dieudonné Mumba Ngoyi, Mari Yamasue, Kosaku Komiya, Kazufumi Hiramatsu, Naoto Uemura, Yuki Sugiura, Mayo Yasugi, Yuka Yamagishi, Hiroshige Mikamo, Satoshi Shiraishi, Takehiro Izumo, Sachie Nakagama, Chihiro Watanabe, Yuko Nitahara, Evariste Tshibangu-Kabamba, Hiroshi Kakeya, Yasutoshi Kido

## Abstract

**Background:** The durability and cross-neutralizability of protective antibodies against evolving SARS-CoV-2 variants are primary concerns in mitigating (re-)exposures. The role of antibody maturation, the process whereby selection of higher avidity antibodies augments host immunity, to determine SARS-CoV-2 neutralizability was investigated.

**Methods:** Sera collected from SARS-CoV-2 convalescent individuals at 2- or 10-months after recovery, and BNT162b2 vaccine recipients at 3 or 25 weeks post-vaccination, were analyzed. Anti-spike IgG avidity was measured on a urea-treated ELISA platform. Neutralizing ability of antibodies was assessed by surrogate virus neutralization. Fold change between variant and wild-type antigen neutralizability was calculated to infer breadth of neutralizability.

**Results:** Compared with early-convalescence, the avidity index of late-convalescent sera was significantly higher (median 37.7 (interquartile range 28.4–45.1) vs. 64.9 (57.5–71.5), p < 0.0001), indicative of progressive antibody maturation extending months beyond acute-phase illness. The urea-resistant, high-avidity fraction of IgG was best predictive of neutralizability (Spearman’s r = 0.49 vs. 0.67 for wild-type; 0.18–0.52 vs. 0.48–0.83 for variants). Higher-avidity convalescent sera showed greater cross-neutralizability against SARS-CoV-2 variants (p < 0.001 for Alpha; p < 0.01 for Delta and Omicron). Vaccinees experienced delayed maturation kinetics, translating to limited breadth of neutralizability at week-25 post-vaccination which was only comparable to that of early-convalescence.

**Conclusions:** Avidity maturation grants broader neutralizability that is resilient against emerging SARS-CoV-2 variants. With immunopotentiation through repeat vaccinations becoming a pivotal strategy to accomplish herd immunity, understanding the variable longitudinal evolutions of the two building blocks of ‘hybrid immunity’ is crucial.

## Introduction

Given the reported coronavirus disease 2019 (COVID-19) re-infections and vaccine breakthrough infections, it is unlikely that convalescent or vaccinated individuals will attain lifelong immune protection [1,2]. Therefore, the magnitude of partial immunity those individuals acquire is an important issue. In the convalescent phase and thereafter, circulating levels of neutralizing IgG that target the SARS-CoV-2 receptor binding domain (RBD) are highly correlated with residual protectivity [3,4]. However, the durability of anti-spike IgGs and their potential cross-neutralizability against rapidly evolving severe acute respiratory syndrome coronavirus 2 (SARS-CoV-2) variants are primary concerns in mitigating future re-exposures [5].

Avidity maturation is the biological process whereby antigen-driven selection of higher affinity antibodies augments the host’s long-lasting protective immunity. Continuous antigen presentation at germinal centers promotes the fine-tuning of the antibody’s complementarity toward the epitope. The kinetics and potential implications of maturing antibody avidity in the context of immune protection against SARS-CoV-2 and its emerging variants of concern (VOCs) remains to be fully elucidated [6–8]. In the present study, we correlated the kinetics of serum antibody avidity maturation, which followed SARS-CoV-2 natural infections, as well as vaccine administrations, to the magnitude and breadth of *in vitro* neutralizability. Our observations provide a logical perspective on the variable durability and resilience of the two building blocks needed for the acquisition of ‘hybrid immunity’.

## Materials and Methods

### Human subjects and sample collection

Total 462 convalescent individuals, who had recovered from COVID-19 [9], were recruited at Osaka Metropolitan University and collaborating institutions preceding the emergence of the SARS-CoV-2 VOCs, comprising a mixed cohort from different regions, where the predominant genotypes belonged to B.1 or B.1.1 lineages [10,11]. Serum samples from recipients of the BNT162b2 SARS-CoV-2 mRNA vaccine were collected at week 3 and week 25 after the first dose. Analyses were conducted in accordance with the 1964 Declaration of Helsinki and its later amendments. This research was approved by the Ethical Committee of Osaka Metropolitan University Graduate School of Medicine (#2020-003). All participants provided written informed consent prior to enrollment.

### Anti-spike IgG measurement

Abbott SARS-CoV-2 IgG II Quant assays (Abbott, IL, USA, 6S6023) were run in accordance with the manufacturer’s instructions. Anti-SARS-CoV-2 IgG enzyme-linked immunosorbent assay (ELISA) (Euroimmun, Lübeck, Germany, EI 2606–9601) was performed in accordance with the manufacturer’s instructions.

### Avidity ELISA

Serum samples were tested for their avidity towards the SARS-CoV-2 spike antigen using an ELISA-based assay (Euroimmun, Lübeck, Germany, EI 2606–9601) [12]. Samples were processed, in an extra pair of wells, with additional urea treatment (5.5 M for 10 min at 37°C) following the incubation of the spike antigen-coated plate with serum samples, to detach any low-avidity antibodies. The avidity index (AI) was calculated as follows: AI = {(OD_450_ of sample with 5.5 M urea) - (OD_450_ of negative control with 5.5 M urea)} / {(OD_450_ of sample without urea) - (OD_450_ of negative control without urea)} × 100 [%]. Serum samples were diluted 1:101 for analysis, except for the post-vaccine sera, which required a higher dilution of 1:201 to bring the OD_450_ signals into linear range.

### Neutralizing ability

Neutralizing ability against wild-type (WT) and VOC SARS-CoV-2 was quantified using the GenScript SARS-CoV-2 sVNT (GenScript, Piscataway, NJ, USA, L00847-A), a competition ELISA-based surrogate virus neutralization test (sVNT). Obtained signals have been shown to strongly correlate with the results of the conventional live-virus neutralization test [13,14]. The assay protocol followed that of the manufacturer’s instructions. Serum samples and controls were pre-incubated to neutralize WT RBD (WT; GenScript, Z03594), or B.1.1.7 (Alpha; GenScript, Z03595), B.1.617.2 (Delta; GenScript, Z03614), or B.1.1.529 (Omicron; GenScript, Z03730) RBD variants conjugated with horseradish peroxidase. The inhibition rate (%inhibition) was calculated as follows: %inhibition = {1 – (OD_450_ of sample) / (OD_450_ of negative control)} × 100 [%].

### Breadth of neutralizability

For quantitative assessment of the serum antibodies’ abilities to cross-neutralize SARS-CoV-2 and its variants, the sVNT %inhibition values were converted to the linearly scaled WHO International Standard units, IU/mL [15,16]. We obtained a standard inhibitory dose-response curve by fitting the log-transformed IU/mL concentration values and their corresponding %inhibition data, obtained from testing four-fold serially diluted WHO International Standard (20/136) in the sVNT, to a four-parameter logistic equation (GraphPad Prism, version 9). The neutralizing titer (IU/mL) of the participants’ serum samples, tested at optimized dilutions (up to 1:30, adjusted to bring the OD_450_ signals into quantitative range of the standard curves), was converted from sVNT %inhibition data by interpolation from the standard curve per different RBD antigen. The fold changes in neutralizing titer between the VOCs and WT (NT_Alpha_/NT_WT_, NT_Delta_/NT_WT_, and NT_Omicron_/NT_WT_) were calculated to serve as the index for the ‘breadth of neutralizability’. Similar indices have served as quantitative measures of relative neutralizing potency towards VOC strains [17].

### Statistical analysis

Differences in titer, avidity, and the magnitude/breadth of neutralizing ability of sera were tested by Mann-Whitney test. For the longitudinal anti-spike IgG titer plot, a linear fit was performed, and Pearson’s correlation coefficient was calculated. Spearman’s correlation coefficient was calculated to assess the strength of the correlation between antibody titer and *in vitro* neutralizability. P-values less than 0.05 were considered statistically significant.

## Results

### Serum anti-SARS-CoV-2 antibodies decay, while maturing in avidity, over time

As recurrently reported, serum levels of circulating anti-spike IgG showed longitudinal decline in titer in SARS-CoV-2 convalescent individuals (Figure 1, panel A, Pearson’s r = −0.28, *p* = 0.049) [18]. To further investigate the qualitative evolution of serum antibodies, representative subsets of sera matched by their total anti-spike IgG titers (Figure 1, panel B, median OD_450_ 1.12 (IQR 0.93–1.25) vs. 1.09 (IQR 0.89–1.26), *p* = 0.98) were selected from early- and late-convalescent (n = 29, median 8 (IQR 7–9) weeks and n = 26, median 37 (IQR 36–40) weeks post-onset of symptoms, respectively) phases, and compared of their serum antibody avidity characteristics. Polyclonal antibodies present in the late-convalescent sera were more avid against the SARS-CoV-2 spike antigen, and thus their OD_450_ were relatively resistant to urea treatment (Figure 1, panel B). The difference in maturity of convalescent sera was reflected in the avidity index (AI), with that of late-convalescent sera demonstrating higher AIs (Figure 1, panel C, median AI 37.7 (IQR 28.4–45.1) vs. 64.9 (IQR 57.5–71.5), *p* < 0.0001), indicating a progressive maturation process that extends months beyond the acute phase of COVID-19.

**Figure 1.**
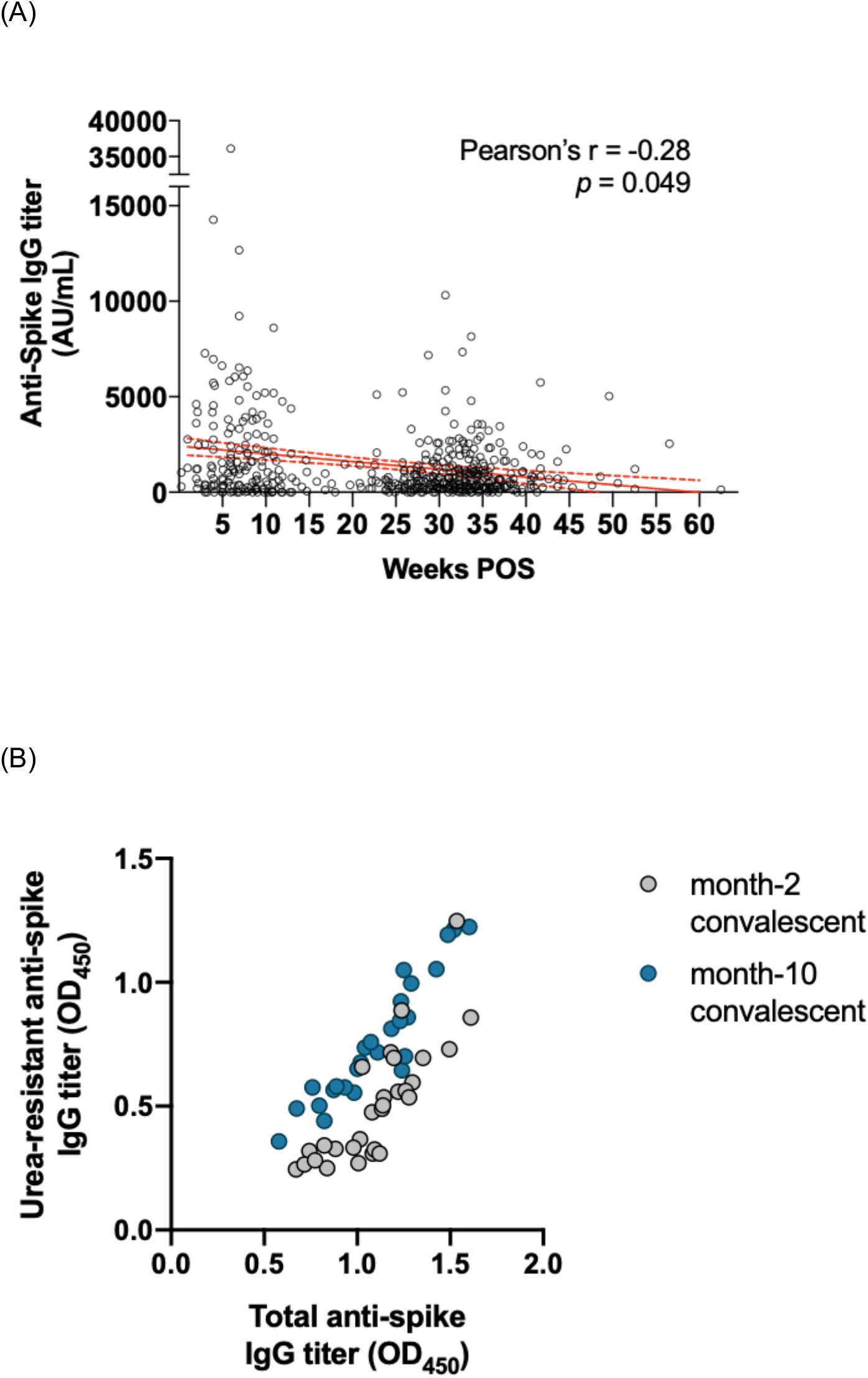
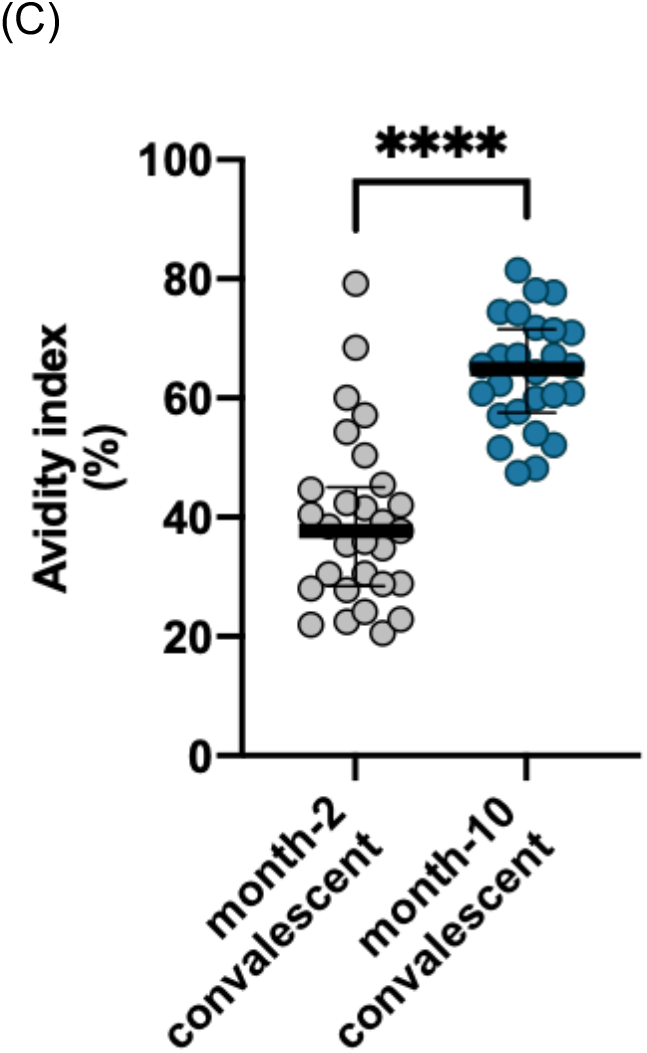
Evolution of anti-SARS-CoV-2 IgG in convalescent individuals. A) Anti-spike IgG levels of individuals observed across the convalescent phase, and their linear fit (solid red line) with 95% confidence intervals (dashed red lines). B) Resistance of the OD_450_ signal against urea treatment in subsets of sera from early- (n = 29) and late-convalescent (n = 26) samples, matched by total anti-spike IgG titers. C) Avidity index of antibodies present in early- (n = 29) and late-convalescent (n = 26) sera. OD_450_, optical density at 450 nm. Statistical significance: \*\*\*\**p* < 0.0001.

### Higher-avidity antibodies exert broadened neutralizability against VOCs

Next, we investigated the functional consequences of enhanced antibody avidity. Compared with the raw OD_450_ values (Figure 2, panel A, upper), the urea-resistant OD_450_ signals from the avidity ELISA (Figure 2, panel A, lower) better correlated with the inhibition rate (%inhibition) obtained from the surrogate sVNT. The results indicated a central role for high-affinity anti-spike IgG antibodies in determining *in vitro* neutralizability against SARS-CoV-2 WT and variant RBDs.

**Figure 2.**
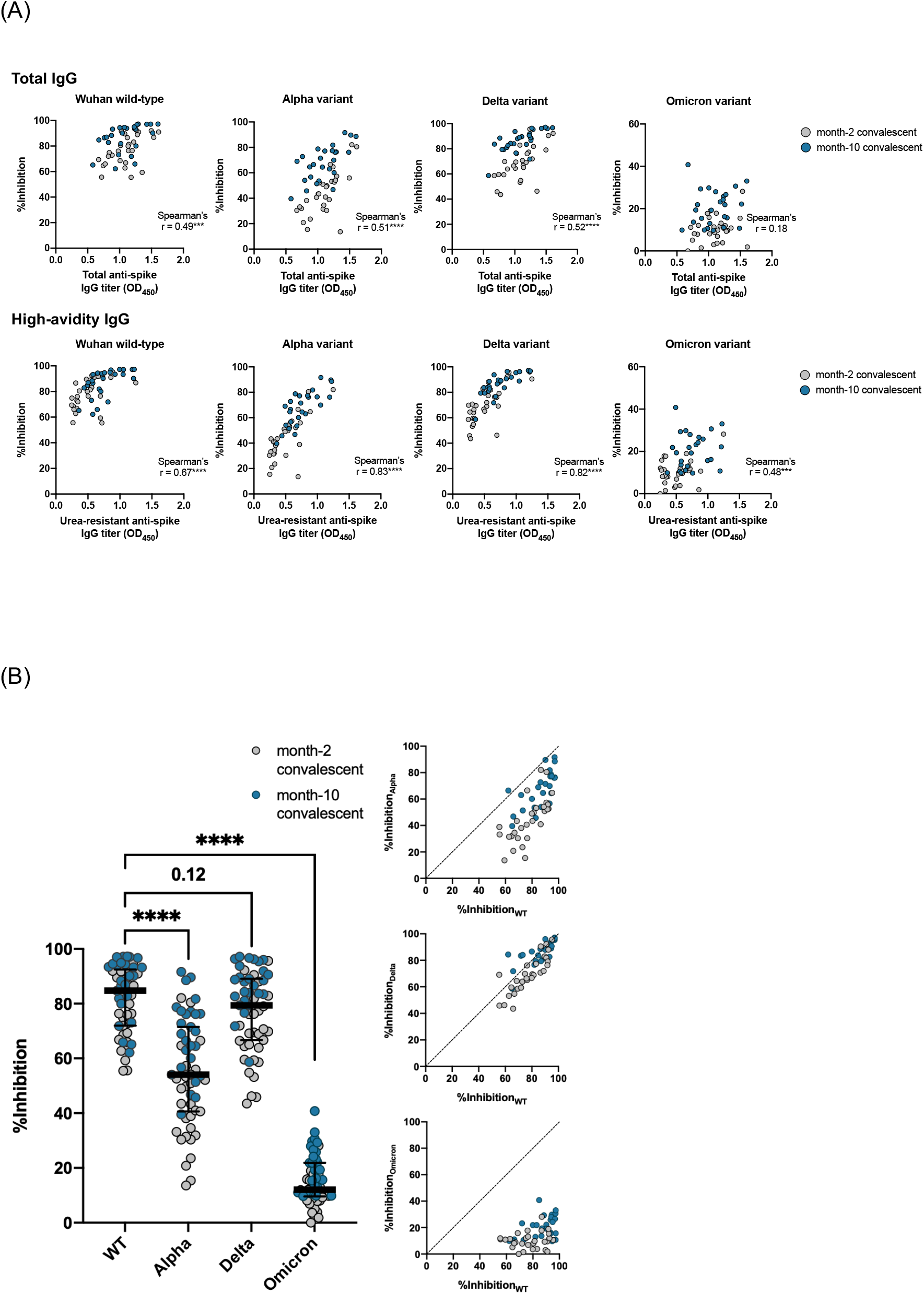
Central role of high-avidity IgG in neutralizing SARS-CoV-2. A) Neutralizability of antibodies present in early- (n = 29) and late-convalescent sera (n = 26) against wild-type and variant SARS-CoV-2, as measured by surrogate virus neutralization, and plotted against total (upper) and urea-resistant, high-avidity anti-spike IgG titers (lower). B) Resistance of SARS-CoV-2 variants against neutralization by convalescent sera. OD_450_, optical density at 450 nm. Statistical significance: \*\*\**p* < 0.001, \*\*\*\**p* < 0.0001.

We next tested the hypothesis that serum antibody avidity determines the tolerability against RBD variations. As previously reported, the Alpha, Delta, and Omicron variants were more resistant to neutralization by convalescent sera, showing attenuated inhibition rates in the sVNT (Figure 2, panel B). To further compare the cross-neutralizability of individual sera against different VOCs, in other words the ‘breadth of neutralizability’, we attempted a linear scale conversion of the neutralizing titer by modifying previously employed methods [15]. Briefly, the neutralizing titer [IU/mL] of each convalescent sample was computed from their %inhibition values [%], through interpolation from standard curves that were generated by testing serially-diluted WHO International Standard (20/136) sera (Figure 3, panel A). The ‘breadth of neutralizability’ was expressed as the NT against the Alpha, Delta, and Omicron VOC RBDs (NT_Alpha_, NT_Delta_, and NT_Omicron_, respectively) relative to the NT against the WT RBD (NT_WT_) in fold changes. Interestingly, when NTs against SARS-CoV-2 VOCs were plotted against that of WT, with respect to the serum antibody’s AI, sera of higher AIs tended to show higher NT_Alpha_, NT_Delta_, and NT_Omicron_ values relative to NT_WT_ (Figure 3, panel B). Next, the fold change in NT against SARS-CoV-2 VOCs relative to the WT was compared between low- (below the 50th percentile) and high-avidity (50th percentile and above) serum samples. Higher-avidity sera showed greater fold change in NT across the different SARS-CoV-2 VOCs (Figure 3, panel C). Altogether, the results indicated that sera of higher AIs exhibited enhanced breadth of neutralizability and better tolerated the variations in SARS-CoV-2 RBD.

**Figure 3.**
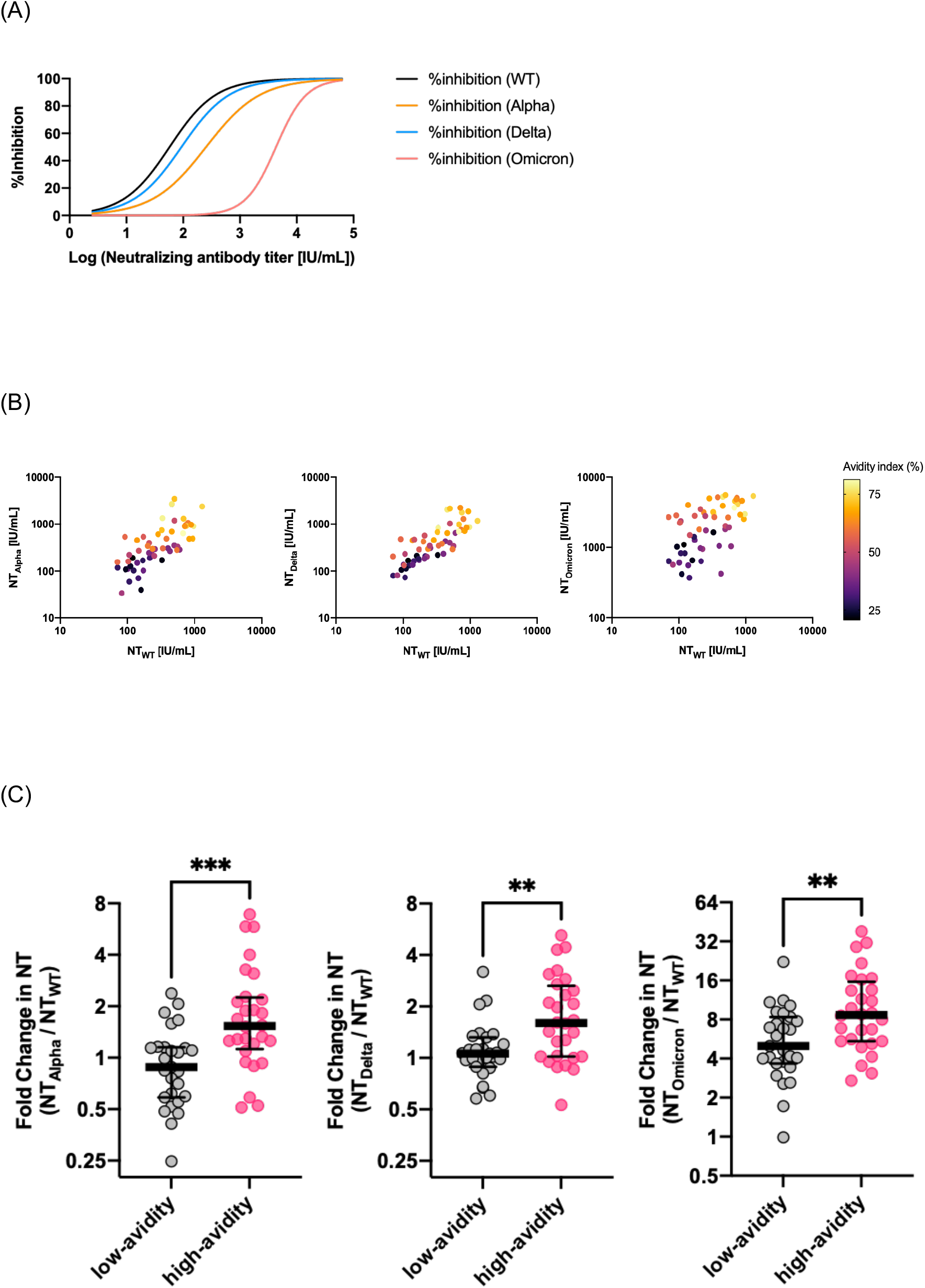
Enhanced neutralization breadth of higher-avidity anti-SARS-CoV-2 IgG. A) Four-parameter logistic standard curves for conversion of inhibition rates to neutralizing antibody titers in WHO International Standard units, IU/mL. B) Cross-neutralizability of convalescent sera against the Alpha (left), Delta (middle), and Omicron variants (right). The samples’ avidity indices are depicted according to the color scale shown. C) Relative inhibition potency towards SARS-CoV-2 variants indexed to the wild-type neutralizing titer, indicating the breadth of neutralizability per serum. Statistical significance: \*\**p* < 0.01, \*\*\**p* < 0.001.

### Vaccinees experience lesser extent of antibody maturation compared with convalescents

The variable kinetics of serum antibody avidity maturation between natural infection immunity and vaccine-elicited immune responses were assessed through analysis of sera from recipients of the BNT162b2 SARS-CoV-2 mRNA vaccine (vaccinees). Sera collected from vaccinees at week 3 (n = 9) and week 25 (n = 8) after the first dose were matched for their total anti-spike IgG titers (median OD_450_ 1.37 (IQR 1.23–1.90) vs. 1.48 (IQR 1.20–1.75), *p* = 1.0), and their AIs were further compared with those of convalescent individuals (Figure 4, panel A). Serum antibodies present in the sera of week-25 vaccinees demonstrated significantly higher AIs compared with those of week-3 vaccinees (median 44.2 (IQR 38.3–47.4) vs 23.4 (IQR 16.9–26.6), *p* = 0.0002), indicating that the second dose of the vaccine had boosted the hosts’ maturation of antibody avidity. However, the AIs of week-25 (approximate month 6) vaccinees were still only equivalent to those of month-2 early-convalescents, depicting a substantial lag in the avidity maturation process unique to the vaccine-elicited immune response (Figure 4, panel B).

**Figure 4.**
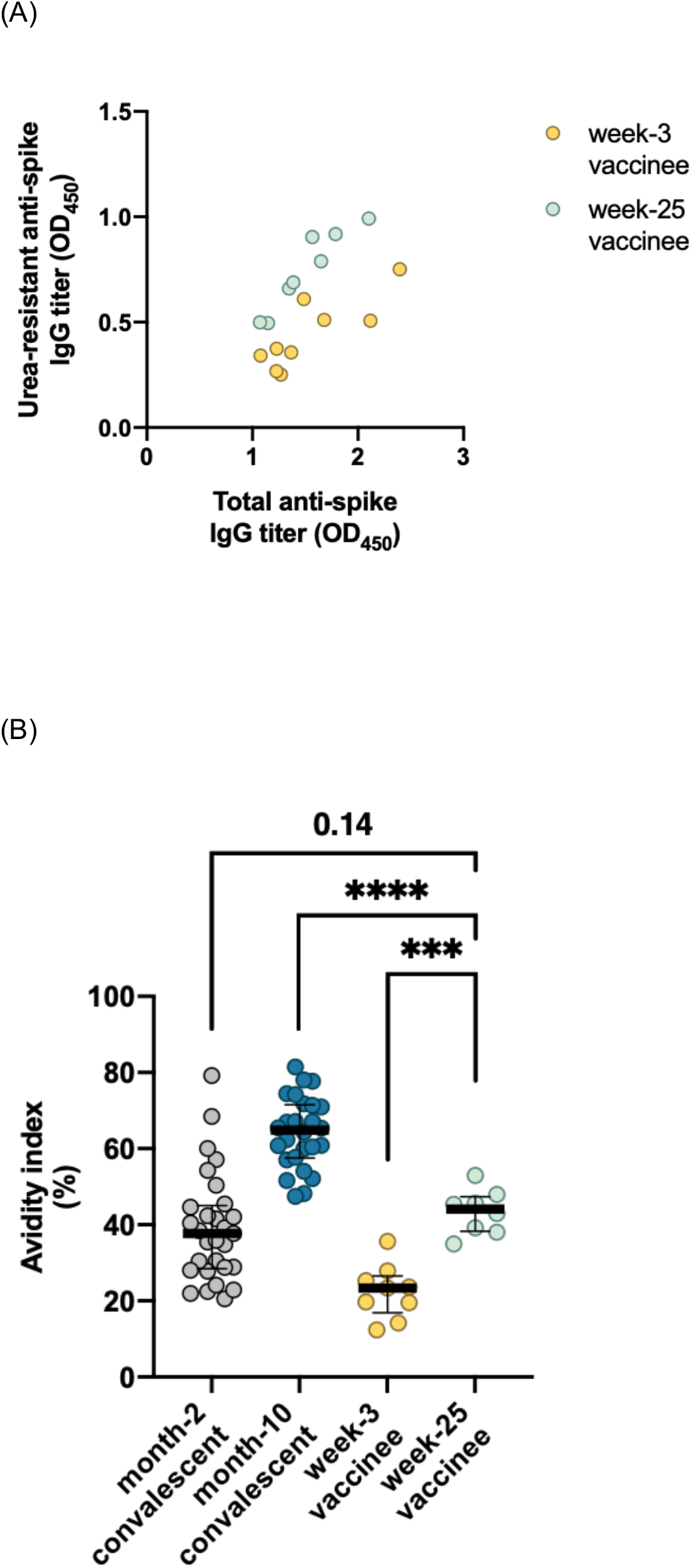
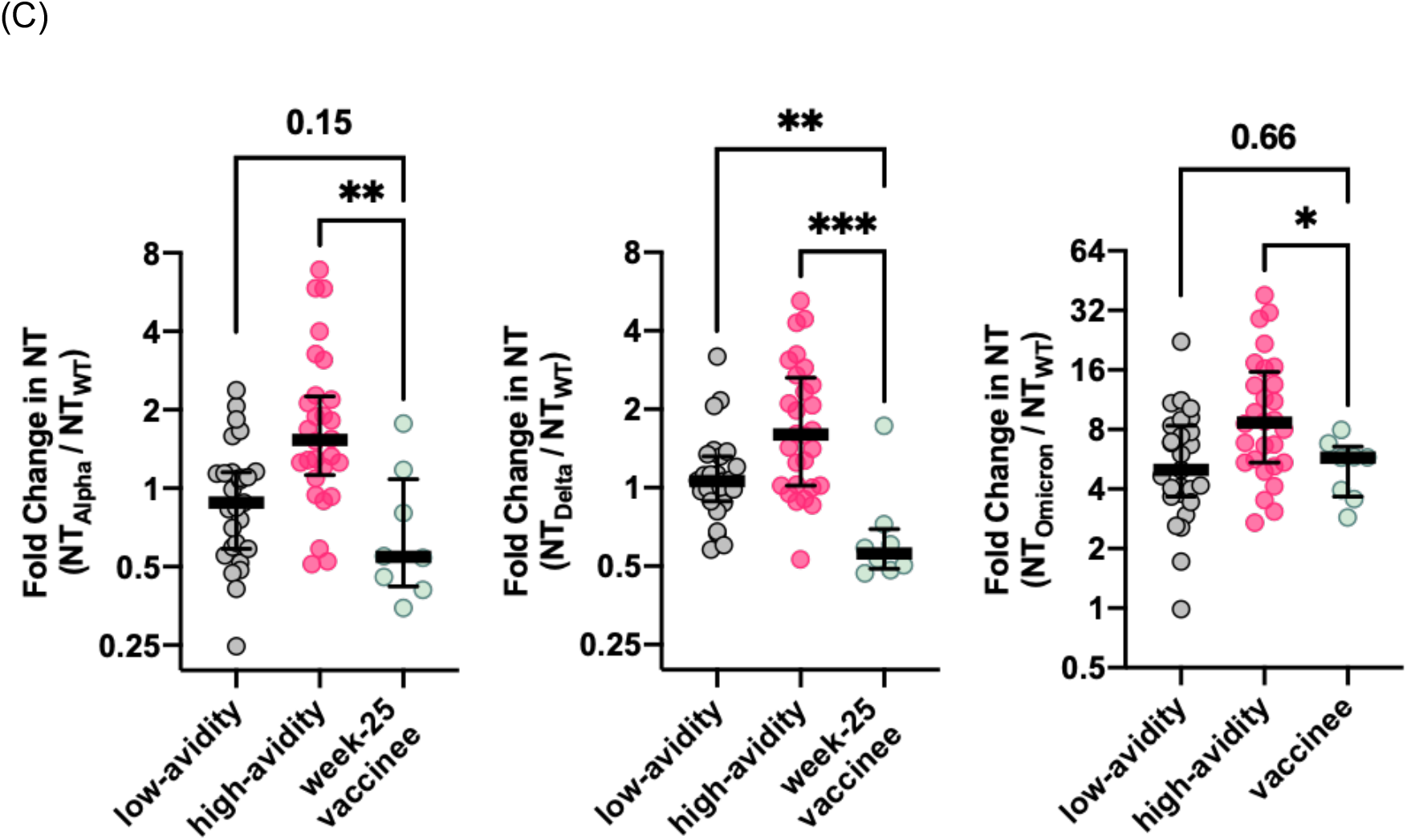
Functional evolution of anti-SARS-CoV-2 IgG in recipients of the BNT162b2 SARS-CoV-2 mRNA vaccine. A) Resistance of the OD_450_ signal against urea treatment in subsets of sera from vaccine recipients at week 3 (n = 9) and week 25 (n = 8) post-vaccination, matched by total anti-spike IgG titers. B) Avidity index of antibodies present in week-3 (n = 9) and week-25 (n = 8) vaccinees in comparison with convalescent individuals. C) Relative inhibition potency of week-25 vaccinee sera (n = 8) towards SARS-CoV-2 variants plotted against that of convalescent individuals. OD_450_, optical density at 450 nm. Statistical significance: \**p* < 0.05, **p < 0.01, ***p < 0.001, ****p < 0.0001.

### Contribution of the matured IgG fraction to neutralizability among vaccinees

Interestingly, in contrast to the convalescent sera and the week-25 vaccinee sera, the neutralizability of the sera of week-3 vaccinees showed a relatively weak correlation (Spearman’s r = 0.52, *p* = 0.16) with their urea-resistant anti-spike IgG titer. The correlation was rather strong (Spearman’s r = 0.87, p < 0.01) when the levels of neutralizability were plotted against total anti-spike IgG titers. This indicates that high-avidity antibodies contribute less to neutralizability early in the vaccine response and become a stronger determinant only after the second dose (Spearman’s r = 0.71, *p* = 0.06). Reflecting their still insufficient degree of maturation, the breadth of neutralizability of the week-25 vaccinees’ sera was limited to an extent comparable to that of early-convalescent sera (Figure 4, panel C).

## Discussions

Antibody avidity maturation in COVID-19 convalescents was observed as an ongoing process that surprisingly extended months beyond the acute phase. This finding supports the notion that SARS-CoV-2 proteins persist in tissues to continuously fuel the germinal centers for antigen presentation, even during late convalescence [19]. More importantly, the high-avidity neutralizing IgGs, which are the result of continuous B-cell evolution, remain in the circulation of late convalescents, fortifying for viral re-encounters by exerting enhanced cross-neutralizability against emerging SARS-COV-2 variants.

In theory, immature plasma cells produce antibodies that possess conformationally flexible antigen-binding surfaces, which at the cost of lowered specificity and affinity, are capable of recognizing diverse antigens (poly-reactivity hypothesis) [20]. In contrast, mature plasma cells produce antibodies of preorganized and rigid paratopes so as to take more energetically favorable configurations with improved complementarity against epitopes [21]. This general tendency towards paratope rigidification upon avidity maturation, while favorable in terms of enhancing specificity, is seemingly detrimental for broad cross-reactivity. In the case of SARS-CoV-2-neutralizing antibodies, however, our results show that high-avidity antibodies exert broader protection against evolving variants, granting long-lasting protective immunity. This paradoxical relationship between affinity maturation and the broadening in reactivity of SARS-CoV-2-neutralizing antibodies is worthy of note. The molecular basis of B-cell evolution is explained by the acquisition of somatic hypermutations [22]. The aforementioned rigidification of paratopes is only one possible consequence of such somatic hypermutations. Mutations may lead to increased buried surface area upon complex formation, strengthening antibody-antigen binding. B-cells may alternatively acquire amino acid substitutions that result in better complementarity of the antigen binding site. Additive molecular contacts at the paratope-epitope interface, through the addition of novel polar or hydrophobic bonds, may also underlie the better complementarity [23]. The increase in variety of the structural determinants shall decrease the dependency of the paratope-epitope interaction on specific amino acid residues [24]. Thus, the latter diverse biophysical mechanisms of antibody affinity maturation all provide explanations to the tolerability of high-affinity antibodies against SARS-CoV-2 variants.

Given that the majority of the world’s population has relied largely on the SARS-CoV-2 vaccines for immunological protection, our secondary interest was how the vaccine-elicited immune responses were different from natural infection-acquired immunity. The kinetics of affinity maturation of SARS-CoV-2-specific antibodies have recently been shown to correlate with the longevity of immune protection following natural SARS-CoV-2 infection [25–27]. The here observed higher potency of the high-avidity antibodies in neutralizing SARS-CoV-2 may counter the longitudinal decay in quantity and explain the durability of infection-acquired immune protection. Interestingly, compared with natural infection, we observed a substantial delay in the maturation process of vaccine-elicited antibodies. This was accompanied by still insufficient cross-neutralizability against SARS-CoV-2 variants even 6 months after vaccination. Large-scale clinical trials in real-world settings have shown greater protection associated with infection-acquired immunity than with vaccine-elicited immune responses [28,29]. The present study further enriches the evidence for and provides an immunological basis to the greater protection associated with infection-acquired immunity. Additionally, the urea-resistant fraction of the antibody titer was predictive of neutralizability only following the second dose of the vaccine. In contrast to the limited variety of epitopes recurrently targeted across convalescent individuals [30], the vaccine-elicited immune response is mapped to a broadened epitope landscape [31,32]. During the initial response to the SARS-CoV-2 vaccine, the majority of urea-resistant signals obtained from our avidity ELISA were possibly from a widely dispersed, non-neutralizing antibody repertoire that still dominated the week-3 vaccinees’ sera. The second vaccine dose seemingly facilitated the positive selection of neutralizing over non-neutralizing epitopes. Collectively, the dynamics of epitope selection, as well as avidity maturation, seem to radically differ among vaccinees and convalescents [33].

Furthermore, circulating IgG may serve as sources of passive immunotherapeutics [34]. In such a context, their extent of maturity may affect cross-neutralizability and immunotherapy outcome. Immunotherapy using convalescent sera has unfortunately remained investigational for COVID-19, though expected beneficial for those with deficient active immunity [35,36]. To mitigate the emergence of immune-escaping variants in future pandemics, assessing avidity to stratify the potency of donated sera may be a reasonable strategy for effective immunotherapy.

Limitations of the present study include: (i) the serum samples were not serial collections from individuals, (ii) the cellular compartment was not studied, which also plays decisive roles in immune protection, and (iii) the observation period of vaccinees was limited, so the long-term consequences remain elusive.

In conclusion, the present study showed that the continuous process of avidity maturation, extending beyond late convalescence, grants broader neutralizability and robust protection that stand resilient against emerging SARS-CoV-2 variants. With immunopotentiation through repeat vaccinations becoming a pivotal strategy to accomplish herd immunity, understanding the longitudinal evolution of vaccine-induced immune responses and the incremental protective effects of booster vaccinations, that are to be also repeated in convalescent individuals (‘hybrid immunity’), remains critical.

## Data Availability

All data produced in the present work are contained in the manuscript.

## Funding

This research was supported by the Japan Agency for Medical Research and Development [grant numbers JP20jk0110021 to Y.N., JP20wm0125003 and JP20he1122001 to Y.K.]; the Japan Society for the Promotion of Science KAKENHI [21K09078 to N.K., 22K15927 to Y.N.]; and the Osaka Metropolitan University Strategic Research Grant [grant number OCU-SRG2021_YR09 to Y.N.]. The authors also received support from the Osaka Metropolitan University Special Reserves Fund for COVID-19 and the Shinya Yamanaka Laboratory COVID-19 Private Fund.

## Acknowledgments

The authors would like to thank all participants of the study and, for technical assistance, Mrs. Mika Oku from Osaka Metropolitan University. Data acquisition was partly performed at the Research Support Platform, Graduate School of Medicine, Osaka Metropolitan University.

## Conflicts of Interest

Y.N. and Y.K. report equity ownership of Quantum Molecular Diagnostics, an Osaka Metropolitan University spinout targeting infectious diseases to develop innovative diagnostics. Y.N. and Y.K. also report receiving financial support outside of this work from Abbott Japan LLC, Japan.

## Notes

### Author Declarations

This research was approved by the Ethical Committee of Osaka Metropolitan University Graduate School of Medicine (#2020-003).

## References

[1] Hacisuleyman E, Hale C, Saito Y, et al. Vaccine breakthrough infections with SARS-CoV-2 variants. N Engl J Med, 2021;384:2212–2218. https://doi.org/10.1056/nejmoa2105000.

[2] To KK-W, Hung IF-N, Ip JD, et al. Coronavirus Disease 2019 (COVID-19) Re-infection by a phylogenetically distinct severe acute respiratory syndrome coronavirus 2 strain confirmed by whole genome sequencing. Clin Infect Dis, 2020;2019:1–6. https://doi.org/10.1093/cid/ciaa1275.

[3] Khoury DS, Cromer D, Reynaldi A, et al. Neutralizing antibody levels are highly predictive of immune protection from symptomatic SARS-CoV-2 infection. Nat Med, 2021;27:1205–1211. https://doi.org/10.1038/s41591-021-01377-8.

[4] Bergwerk M, Gonen T, Lustig Y, et al. Covid-19 breakthrough infections in vaccinated health care workers. N Engl J Med, 2021;385:1474–1484. https://doi.org/10.1056/nejmoa2109072.

[5] Cromer D, Juno JA, Khoury D, et al. Prospects for durable immune control of SARS-CoV-2 and prevention of reinfection. Nat Rev Immunol, 2021;21:395–404. https://doi.org/10.1038/s41577-021-00550-x.

[6] Bauer G. The potential significance of high avidity immunoglobulin G (IgG) for protective immunity towards SARS-CoV-2. Int J Infect Dis, 2021;106:61–64. https://doi.org/10.1016/j.ijid.2021.01.061.

[7] Bauer G, Struck F, Schreiner P, Staschik E, Soutschek E, Motz M. The challenge of avidity determination in SARS-CoV-2 serology. J Med Virol, 2021;93:3092–3104. https://doi.org/10.1002/jmv.26863.

[8] Nakagama Y, Nitahara Y, Kaku N, Tshibangu-Kabamba E, Kido Y. A dual-antigen SARS-CoV-2 serological assay reflects antibody avidity. J Clin Microbiol, 2022;60:e0226221. https://doi.org/10.1128/JCM.02262-21.

[9] Adachi T, Ayusawa M, Ujiie M, et al. Novel coronavirus infection COVID-19 medical practice guidelines. Version 7.2. Available at: https://www.mhlw.go.jp/content/000936623.pdf (Accessed 1 July 2022).

[10] Nakagama Y, Komase Y, Candray K, et al. Serological testing reveals the hidden COVID-19 burden among health care workers experiencing a SARS-CoV-2 nosocomial outbreak. Microbiol Spectr, 2021;9:e0108221. https://doi.org/10.1128/Spectrum.01082-21.

[11] Nakagama Y, Rodriguez-Funes MV, Dominguez R, et al. Cumulative seroprevalence among healthcare workers after the first wave of the COVID-19 pandemic in El Salvador, Central America. Clin Microbiol Infect, 2022;S1198-743X(22)00333-0. doi: 10.1016/j.cmi.2022.06.020.

[12] Pichler D, Baumgartner M, Kimpel J, et al. Marked increase in avidity of SARS-CoV-2 antibodies 7–8 months after infection is not diminished in old age. J Infect Dis, 2021:1–7. https://doi.org/10.1093/infdis/jiab300.

[13] Tan CW, Chia WN, Qin X, et al. A SARS-CoV-2 surrogate virus neutralization test based on antibody-mediated blockage of ACE2–spike protein–protein interaction. Nat Biotechnol, 2020;38:1073–1078. https://doi.org/10.1038/s41587-020-0631-z.

[14] Mariën J, Michiels J, Heyndrickx L, et al. Evaluation of a surrogate virus neutralization test for high-throughput serosurveillance of SARS-CoV-2. J Virol Methods, 2021;297:114228. https://doi.org/10.1016/j.jviromet.2021.114228.

[15] Taylor SC, Hurst B, Martiszus I, et al. Semi-quantitative, high throughput analysis of SARS-CoV-2 neutralizing antibodies: Measuring the level and duration of immune response antibodies post infection/vaccination. Vaccine, 2021;39:5688–5698. https://doi.org/10.1016/j.vaccine.2021.07.098.

[16] National Institute for Biological Standards and Control. First WHO International Standard for anti-SARS-CoV-2 immunoglobulin, human. (NIBSC code: 20/136) Available at: https://www.nibsc.org/documents/ifu/20-136.pdf (Accessed 1 July 2022).

[17] Moriyama S, Adachi Y, Sato T, et al. Temporal maturation of neutralizing antibodies in COVID-19 convalescent individuals improves potency and breadth to circulating SARS-CoV-2 variants. Immunity, 2021;54:1841-1852.e4. https://doi.org/10.1016/j.immuni.2021.06.015.

[18] Gaebler C, Wang Z, Lorenzi JCC, et al. Evolution of antibody immunity to SARS-CoV-2. Nature, 2021;591:639–644. https://doi.org/10.1038/s41586-021-03207-w.

[19] Gaebler C, Wang Z, Lorenzi JCC, et al. Evolution of antibody immunity to SARS-CoV-2. Nature, 2021;591:639–644. https://doi.org/10.1038/s41586-021-03207-w.

[20] Furukawa K, Akasako-Furukawa A, Shirai H, Nakamura H, Azuma T. Junctional amino acids determine the maturation pathway of an antibody. Immunity, 1999;11:329–338. https://doi.org/10.1016/S1074-7613(00)80108-9.

[21] Manivel V, Sahoo NC, Salunke DM, Rao KVS. Maturation of an antibody response is governed by modulations in flexibility of the antigen-combining site. Immunity, 2000;13:611–620. https://doi.org/10.1016/S1074-7613(00)00061-3

[22] Victora GD, Nussenzweig MC. Germinal centers. Annu Rev Immunol, 2012;30:429–457. https://doi.org/10.1146/annurev-immunol-020711-075032

[23] Mishra AK, Mariuzza RA. Insights into the structural basis of antibody affinity maturation from next-generation sequencing. Front Immunol, 2018;9:117. https://doi.org/10.3389/fimmu.2018.00117.

[24] Muecksch F, Weisblum Y, Barnes CO, et al. Affinity maturation of SARS-CoV-2 neutralizing antibodies confers potency, breadth, and resilience to viral escape mutations. Immunity, 2021;54:1853-1868.e7. https://doi.org/10.1016/j.immuni.2021.07.008.

[25] Luo YR, Chakraborty I, Yun C, Wu AHB, Lynch KL. Kinetics of severe acute respiratory syndrome coronavirus 2 (SARS-CoV-2) antibody avidity maturation and association with disease severity. Clin Infect Dis, 2021;73:e3095–e3097. https://doi.org/10.1093/cid/ciaa1389.

[26] Löfström E, Eringfält A, Kötz A, et al. Dynamics of IgG-avidity and antibody levels after Covid-19. J Clin Virol, 2021;144:104986. https://doi.org/10.1016/j.jcv.2021.104986.

[27] Chia WN, Zhu F, Ong SWX, et al. Dynamics of SARS-CoV-2 neutralising antibody responses and duration of immunity: a longitudinal study. Lancet Microbe, 2021;2:e240–e249. https://doi.org/10.1016/S2666-5247(21)00025-2.

[28] Gazit S, Shlezinger R, Perez G, et al. Severe acute respiratory syndrome coronavirus 2 (SARS-CoV-2) naturally acquired immunity versus vaccine-induced immunity, reinfections versus breakthrough infections: a retrospective cohort study. Clin Infect Dis, 2022;2:1–7. https://doi.org/10.1093/cid/ciac262.

[29] Hall V, Foulkes S, Insalata F, et al. Protection against SARS-CoV-2 after Covid-19 vaccination and previous infection. N Engl J Med, 2022;386:1207–1220. https://doi.org/10.1056/nejmoa2118691.

[30] Tong P, Gautam A, Windsor IW, et al. Memory B cell repertoire for recognition of evolving SARS-CoV-2 spike. Cell, 2021:1–12. https://doi.org/10.1016/j.cell.2021.07.025.

[31] Nitahara Y, Nakagama Y, Kaku N, et al. High-resolution linear epitope mapping of the receptor binding domain of SARS-CoV-2 spike protein in COVID-19 mRNA vaccine recipients. Microbiol Spectr, 2021;9 e0096521. https://doi.org/10.1128/spectrum.00965-21.

[32] Greaney AJ, Loes AN, Gentles LE, et al. Antibodies elicited by mRNA-1273 vaccination bind more broadly to the receptor binding domain than do those from SARS-CoV-2 infection. Sci Transl Med, 2021;13:1–13. https://doi.org/10.1126/scitranslmed.abi9915.

[33] Wang Z, Muecksch F, Schaefer-Babajew D, et al. Naturally enhanced neutralizing breadth against SARS-CoV-2 one year after infection. Nature, 2021;595:426–431. https://doi.org/10.1038/s41586-021-03696-9.

[34] Tang J, Lee Y, Ravichandran S, et al. Epitope diversity of SARS-CoV-2 hyperimmune intravenous human immunoglobulins and neutralization of variants of concern. iScience, 2021;24:103006. https://doi.org/10.1016/j.isci.2021.103006.

[35] Takakuwa T, Nakagama Y, Yasugi M, et al. Discrepant antigen-specific antibody responses causing SARS-CoV-2 persistence in a patient receiving B-cell-targeted therapy with rituximab. Intern Med, 2021;60:3827–3831. https://doi.org/10.2169/internalmedicine.7884-21.

[36] Takita M, Yoshida T, Tsuchida T, et al. Low SARS-CoV-2 antibody titers may be associated with poor clinical outcomes for patients with severe COVID-19. Sci Rep, 2022;12:9147. https://doi.org/10.1038/s41598-022-12834-w.

